# Elevated RDW is Associated with Increased Mortality Risk in COVID-19

**DOI:** 10.1101/2020.05.05.20091702

**Authors:** Brody H. Foy, Jonathan C.T. Carlson, Erik Reinertsen, Raimon Padros Valls, Roger Pallares Lopez, Eric Palanques-Tost, Christopher Mow, M. Brandon Westover, Aaron D. Aguirre, John M. Higgins

## Abstract

**Background:** Coronavirus disease 2019 (COVID-19) is an acute respiratory illness with a high rate of hospitalization and mortality. Prognostic biomarkers are urgently needed. Red blood cell distribution width (RDW), a component of complete blood counts that reflects cellular volume variation, has been shown to be associated with elevated risk for morbidity and mortality in a wide range of diseases.

**Methods:** We retrospectively studied the relationship between RDW and COVID-19 mortality risk for 1198 adult patients diagnosed with SARS-CoV-2 at 4 Partners Healthcare Network Hospitals between March 4, 2020, and April 28, 2020.

**Results:** Elevated RDW (> 14.5%) was associated with increased mortality in patients of all ages with a risk ratio of 2.5 (95% CI, 2.3 – 2.8). Stratified by age, the risk ratio was 6.2 (4.4 – 7.9, N = 312) < 50 years, 3.2 (2.5 – 4.1, N = 230) 50-60, 2.3 (1.6 – 3.1, N = 236) 60-70, 1.2 (0.7 – 1.8, N = 203) 70-80, and 1.9 (1.5 – 2.3, N = 216) > 80 years. RDW was significantly associated with mortality in Cox proportional hazards models adjusted for age, D-Dimer, absolute lymphocyte count, and common comorbidities (p < 1e-4 for RDW in all cases). Patients whose RDW increased during admission had a ~3-fold elevation in mortality risk compared to those whose RDW did not change.

**Conclusions:** Elevated RDW at diagnosis and an increase in RDW during admission are both associated with increased mortality risk for adult COVID-19 patients at a large academic medical center network.

## Background

Coronavirus disease 2019 (COVID-19) is an acute respiratory illness caused by infection with severe acute respiratory syndrome coronavirus 2 (SARS-CoV-2). COVID-19 has a high rate of hospitalization, critical care requirement, and mortality.^1,2^ Identifying patients at highest risk of severe disease is important for enabling earlier aggressive intervention and for managing local hospital resources to mitigate critical care crises that have affected some hospital systems. On admission, COVID-19 is associated with lymphopenia, occasional thrombocytopenia, and overall leukopenia.^3^ The clinical course for hospitalized patients varies dramatically, with early evidence showing that ICU admission and mortality are associated with elevated D-Dimer and decreasing lymphocyte count.^1,4^ Additional prognostic markers routinely available at the time of admission are urgently needed.

The red blood cell distribution width (RDW) is a standard component of routine complete blood counts (CBC). RDW quantifies the variation of individual red blood cell (RBC) volumes which vary from one cell to the next and for the same cell over as it circulates during its ~105-day lifespan.^5-7^ Elevated RDW is associated with increased risk for a remarkably wide range of morbidity and mortality: all-cause mortality, mortality from heart disease, pulmonary disease, sepsis, influenza, and cancer; complications in heart failure, severity of coronary artery disease and viral hepatitis; advanced stage and grade for many cancers; development of diabetes, chronic obstructive pulmonary disease, stroke, anemia, and many more.^8-17^ RDW thus appears to represent a non-specific marker of illness with the potential to provide general quantitative prognostic value that may be particularly powerful for a new and unknown disease. RDW is the coefficient of variation in RBC volume, or the standard deviation divided by the mean. An increase in RDW must therefore correspond to a decrease in mean RBC volume (MCV), an increase in RBC volume variance, or both. Prior studies have found evidence in some specific conditions that RDW elevation is caused by delayed clearance of older RBCs. Because RBCs characteristically decrease in cellular volume across their lifespan, persistence of these older, smaller cells thus increases volume variance, and this clearance delay coincides with and offsets a net decrease in RBC production.^6,8,18^ These reports thus suggest the possibility that an elevated RDW in some circumstances may reflect a clinical state in which RBC production and turnover have slowed in the setting of increased production and turnover of leukocytes or platelets, such as would occur in inflammation. While a definitive mechanism for RDW elevation is far from established, there is clear evidence that RDW can provide robust risk-stratification among patients diagnosed with the same acute illness. Here we investigated the association between elevated RDW and risk of mortality in COVID-19.

## Methods

### Subjects and study design

Clinical data was retrospectively analyzed for all patients who tested positive for SARS-CoV-2 between March 4, 2020 and April 28, 2020 at one of four Partners Healthcare Network hospitals (N = 6376). Patients who were not subsequently admitted to one of these hospitals were excluded, as were patients who had not been discharged by the study end date. To allow sufficient time for symptom progression, patients with an initial COVID-19 diagnosis occurring after April 21, 2020 were also excluded. The key results described below were similar when using different exclusion windows, and when including current inpatients and outpatients (see **supplemental material**). Inpatients with a total hospital stay less than 24 hours were excluded if discharged alive. These exclusions led to a total cohort of 1198 patients across four medical centers: Massachusetts General Hospital (MGH) 595; Brigham and Women’s Hospital (BWH) 392; North Shore Medical Center (NSMC) 115; Newton-Wellesley Hospital (NWH) 96. Patients with multiple separate inpatient visits related to COVID-19 were treated as having been admitted during the first visit and discharged during the final visit. For analysis purposes, patients who had visits spanning multiple medical centers were classified as being in the cohort associated with the first medical center they visited.

For all inpatients, red blood cell distribution width (RDW), absolute lymphocyte count (lymph), and D-Dimer were collected approximately daily, as part of standard clinical care along with other clinical laboratory tests. RDW and lymphocyte counts were performed on Sysmex XN-9000 automated hematology instruments. D-Dimer was measured on a bioMerieux Vidas 3 instrument. SARS-CoV-2 was diagnosed using multiple instruments and assays including bioMerieux BioFire, Roche Cobas 6800, and Cepheid GeneXpert. Comorbidities were analyzed by identifying ICD10 codes associated with each patient in their diagnostic history. Mortality was determined by reviewing discharge summaries, with an assumption of no COVID-19-related deaths for patients who were discharged alive. The effect of this assumption is explored in the **supplemental material**.

Within the main text, results are presented using pooled data from MGH, BWH, NSMC and NWH. Qualitatively and quantitatively similar results are seen when analyzing cohorts from BWH and MGH separately (see **supplemental material;** NSMC and NWH are not analyzed separately due to smaller cohort numbers). All patient data was gathered using the Partners Healthcare Research Patient Data Registry (RPDR) and Electronic Data Warehouse (EDW), under a research protocol approved for a waiver of consent by the Partners Healthcare Institutional Review Board.

### Statistical analysis

The Kaplan-Meier method was used to analyze survival in inpatients stratified by RDW at admission. To account for age as a confounder, patients were grouped into 5 categories: <50yrs, 50-60yrs, 60-70yrs, 70-80yrs, and 80+ yrs old. An abnormal RDW was defined as >14.5%, the current upper limit of the healthy adult reference interval at both MGH and BWH. Patients who were discharged alive were censored on April 28, 2020, with results for other censoring choices presented in the **supplemental material**.

Mortality hazard ratios were calculated using a Cox proportional-hazards model. Models were fit with univariate inputs and multivariate inputs, using RDW and three known risk factors for COVID-19 prognosis: age, absolute lymphocyte count, and D-Dimer.^4^ Models were also fit treating variables as either continuous or binarized with a risk threshold. Risk thresholds were defined as age > 70yrs, RDW > 14.5%, lymph < 0.8, and D-Dimer < 1500. Thresholds for age, lymph, and D-Dimer were chosen based on prior COVID-19 reports.^4^ All thresholds produced similar percentages of high-risk patients within the cohort (34%, 30%, 26%, 26% for age, RDW, lymph and D-Dimer respectively). For continuous models, hazard ratios were normalized based on meaningful clinical changes in the measurement: 10yrs for age, 0.5% for RDW, 0.1 ×10^9^/L for lymph, and 100 mg/L for D-Dimer. For ease of comparison, hazard ratios for lymphocyte count were inverted, to represent the increased hazard for a decrease in value. All other ratios were relative to increases. Multivariate proportional-hazards models were also fit using patient comorbidities, with results presented in the **supplemental material**.

Changes in RDW over hospital stay were evaluated by taking the difference between the first and last available RDW measurement. RDW trajectories were plotted for patients stratified by normal and abnormal RDW, as well as survival status on discharge. Mean RDW trajectories were calculated by linearly interpolating patient RDW values and calculating the mean value for all patients in a cohort at the interpolated time post admission (curves were calculated using a temporal spacing of 1hr). Average RDW trajectories were calculated over the first week of admission, and cohorts were limited to patients who had a hospital stay of at least 7 days.

Statistical differences between incidence rates were analyzed using a Chi-square proportion comparison test. All statistical analysis was performed in MATLAB 2019b, using the Statistical Analysis Toolbox (MathWorks, Natick MA).

## Results

### Baseline characteristics of patients

We retrospectively investigated the association between RDW measured at the time of admission for diagnosis of SARS-CoV-2 infection and the risk of mortality in 1198 adult patients between March 4, 2020, and April 28, 2020. Baseline characteristics of patients are shown in Table 1.

**Table 1.** **Patient Characteristics**. All data are presented either as percentages, or as mean (std), *except for D-Dimer, which is presented as median (25th-75th), due to its long upper tail.

| Demographics | Survivors | Non-Survivors |
| --- | --- | --- |
| N | 1043 | 155 |
| Age - yrs | 59.6 (17.5) | 75 (13.2) |
| Sex - % male | 54% | 63% |
| BMI - kg/m<sup>2</sup> | 30.7 (7.3) | 30.4 (7.1) |
| **RDW - %** |  |  |
| Age < 50 | 13.6 (2.0) | 15.6 (3.2) |
| Age: 50-60 | 13.6 (1.7) | 15.9 (3.3) |
| Age 60-70 | 14.1 (1.9) | 15.3(2.4) |
| Age 70-80 | 14.3 (2.0) | 14.6 (1.9) |
| Age 80+ | 14.4 (1.9) | 15.2 (1.9) |
| Entire cohort | 13.9 (1.9) | 15.2 (2.3) |
| **Other Laboratory Tests** |  |  |
| Absolute lymphocyte count - N x 10<sup>9</sup>/L | 1.25 (1.33) | 0.91 (0.7) |
| D-Dimer - mg/L | 866 (523-1652) | 1499 (875-2368)* |
| Hematocrit - % | 37.7 (6.1) | 35.3 (7.3) |
| Hemoglobin -g/dL | 12.4 (2.2) | 11.4 (2.5) |
| Mean corpuscular hemoglobin - pg | 29 (2.5) | 29.2 (2.8) |
| Mean corpuscular hemoglobin concentration - g/dL | 32.8 (1.5) | 32.3 (1.7) |
| Platelet count - 10<sup>3</sup>/μL | 236 (108) | 193.2 (108) |
| Red blood cell count - $10^6/\mu\text{L}$ | 4.3 (0.8) | 3.9 (0.9) |
| White blood cell count - $10^3/\mu\text{L}$ | 7.5 (8.5) | 9.2 (5.7) |
| **Other Outcomes** |  |  |
| Length of hospital stay - days | 5.4 (5.6) | 5 (5.7) |
| **Comorbidities - %** |  |  |
| Any | 36% | 55% |
| COPD | 4% | 10% |
| Diabetes Mellitus | 16% | 23% |
| Hypertension | 22% | 40% |
| Coronary Artery Disease | 8% | 18% |
| Chronic kidney disease | 9% | 23% |

### Elevated RDW at admission for COVID-19 is associated with increased mortality risk

Patients whose RDW was above 14.5% at admission had a 22% mortality risk, while those with RDW <= 14.5 had a mortality risk of 8.7%. The relative risk of mortality for those with elevated RDW was 2.5 (2.25 - 2.83). See Figure 1 and Table 2. Age has previously been shown to be a strong risk factor for COVID-19 mortality.^4^ In patient groups stratified by age, elevated RDW remained associated with increased relative risk of mortality for patients < 50, 50-60, 60-70, and 80+ yrs. For patients 70-80yrs, the risk ratio (1.2) was not statistically distinguishable from 1.0. For younger patients (< 70yrs), the relative risk associated with elevated RDW was particularly high within 48 hours of admission. Patients 50-70yrs with elevated RDW at admission in our cohort experienced a mortality rate of 9.1% (12/131) within 48 hours of admission, while the rate for those with RDW <= 14.5 was 2.1% (7/335).

**Figure 1.**
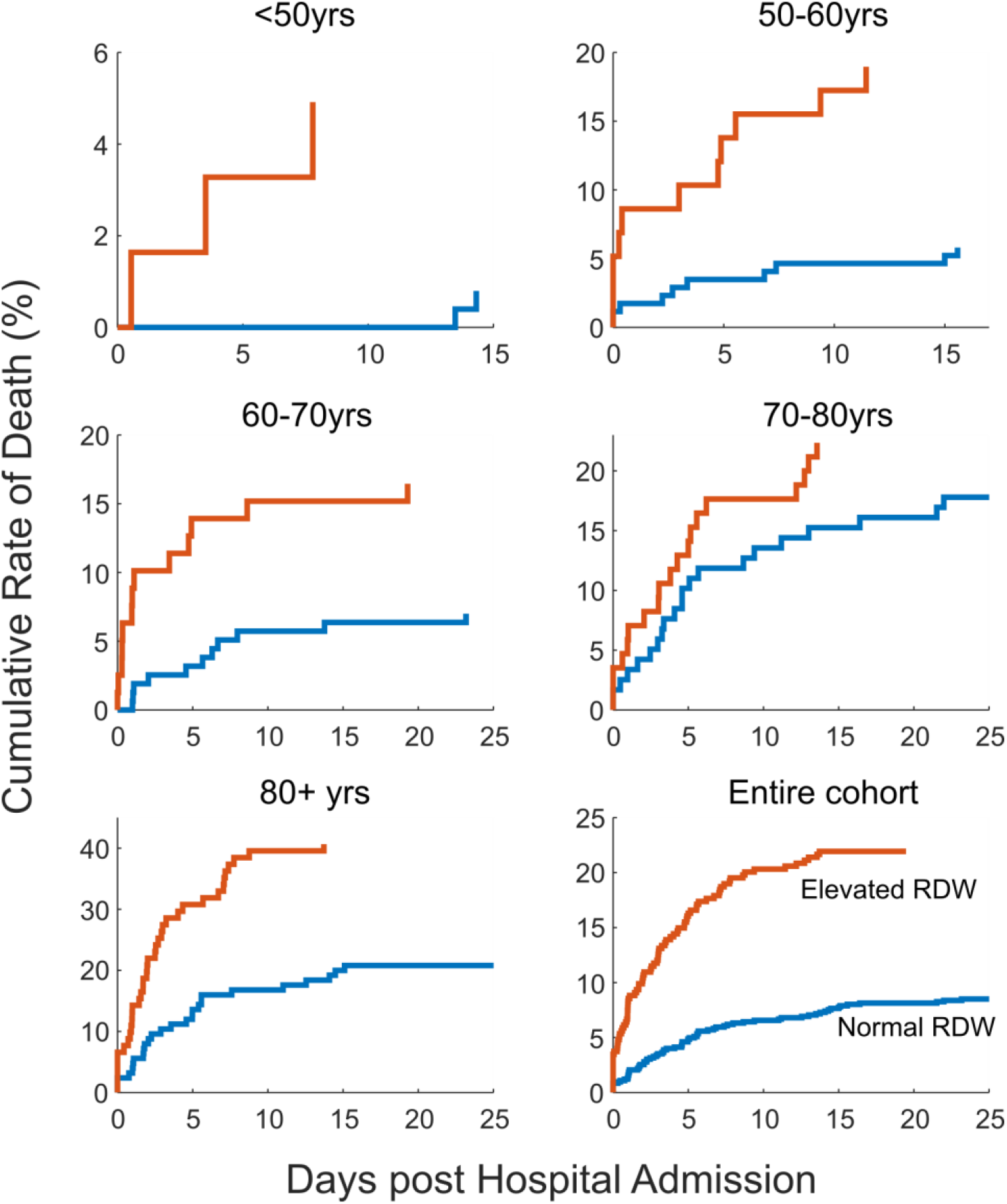
**Elevated RDW (> 14.5%) at hospital admission is associated with increased mortality risk among COVID-19 patients**. Across all adult ages, RDW > 14.5% measured at the time of admission is associated with a 22% mortality rate, compared to an 8.7% for patients whose RDW at admission was within the reference interval. All increases are statistically significant except for 70-80 years. RDW-stratified mortality rates are highly divergent in our study cohort for the 50-60 and 60-70 year age ranges soon after admission. See Table 2 for age- and RDW-stratified mortality rates.

**Table 2.** Mortality rates stratified by age and RDW elevation (> 14.5%) at admission.

|  | Normal RDW |  | Elevated RDW |  |  |  |
| --- | --- | --- | --- | --- | --- | --- |
| Age | N | Mortality | N | Mortality | p-value | Risk Ratio (95% confidence interval) |
| <50yrs | 251 | 0.8% | 61 | 4.9% | 0.02 | 6.17 (4.4-7.94) |
| 50-60yrs | 172 | 5.8% | 58 | 19.0% | 0.003 | 3.26 (2.46-4.07) |
| 60-70yrs | 157 | 7.0% | 79 | 16.5% | 0.02 | 2.35 (1.59-3.1) |
| 70-80yrs | 118 | 18.6% | 85 | 22.4% | 0.52 | 1.2 (0.65-1.75) |
| 80+ yrs | 125 | 21.6% | 91 | 40.7% | 0.002 | 1.88 (1.47-2.3) |
| Entire cohort | 824 | 8.7% | 374 | 22.2% | 1.35E-10 | 2.54 (2.25-2.83) |

### RDW-associated risk is independent of D-Dimer and Lymphopenia

Prior studies have found elevated D-Dimer and low absolute lymphocyte count to be associated with increased mortality risk.^4^ We performed Cox proportional hazards regression modeling to investigate whether RDW provided additional predictive information beyond these markers, both when considered as a binary marker relative to the 14.5% reference interval boundary and when considered as a continuous marker. Figure 2 shows that RDW had a significant effect on risk for all models considered, including those adjusting for age, lymphocyte count, and D-Dimer levels, both as continuous and binarized predictors. In the **supplemental material** we performed further Cox proportional hazards modeling, incorporating five major comorbidities (chronic obstructive pulmonary disease, coronary artery disease, chronic kidney disease, diabetes mellitus, and hypertension). When jointly modeled, the risk ratio associated with RDW > 14.5% remained significant and was larger than that for any comorbidity (RDW risk ratio: 1.87 (1.35-2.6), p-value < 0.001).

**Figure 2.**
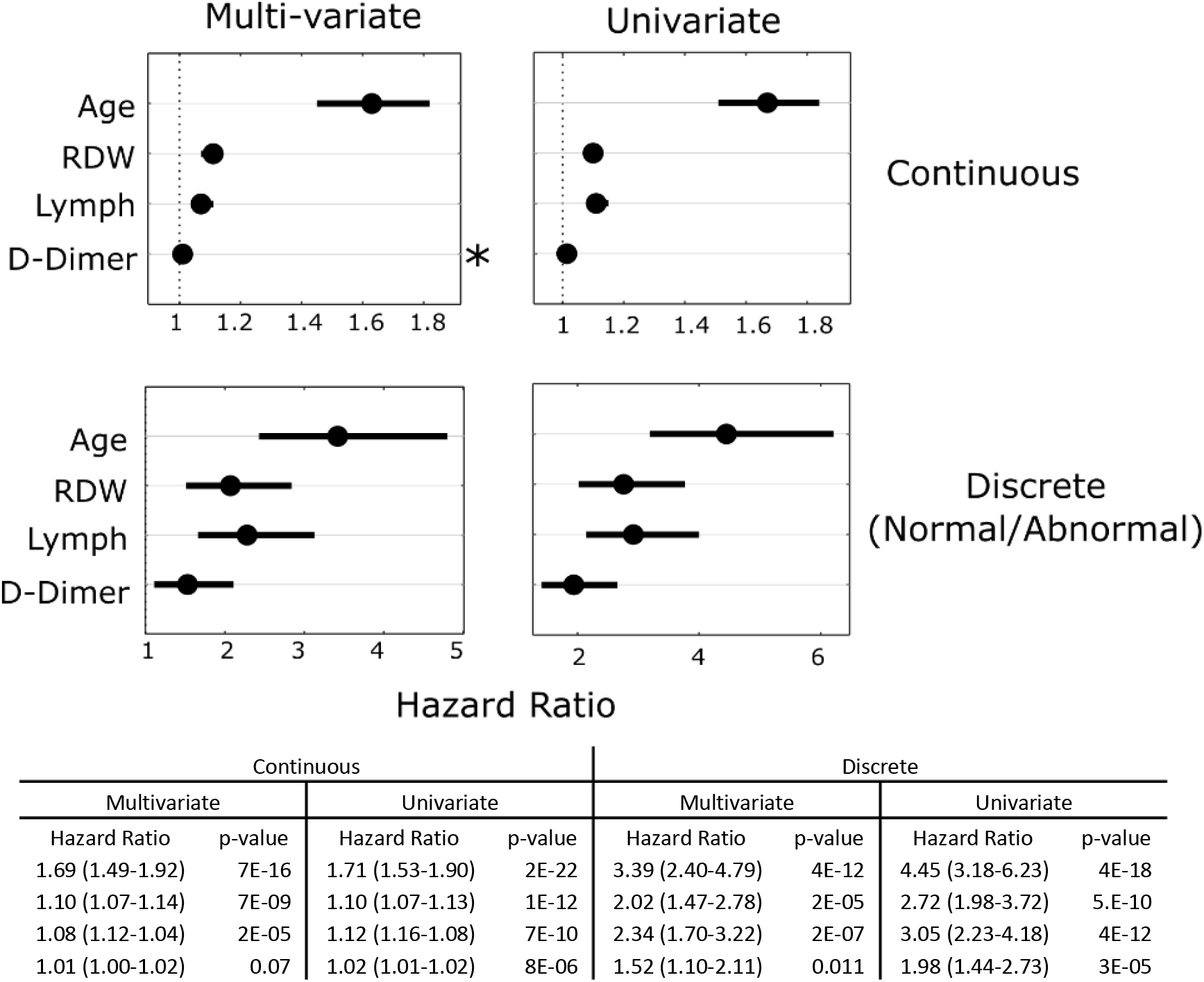
**Cox proportional hazards modeling of mortality risk finds RDW associated with increased mortality risk after adjusting for age, absolute lymphocyte count, and D-Dimer**. The top row shows results from models of mortality as a function of age, RDW, lymphocyte count, and d-dimer treated as continuous variables, in a multivariate model (top left) and each variable separately considered in a univariate model. Variables were normalized as follows: age to an increase of 10 years, RDW increase of 0.5%, D-Dimer increase of 100 mg/L, and a lymphocyte decrease of 0.1 103/ul. The bottom row shows results for models using discrete variables with the following thresholds: age > 70yrs, RDW> 14.5%, lymph < 0.8, and D-Dimer > 1500. These thresholds provide a similar fraction of abnormality in the cohort (33%, 29%, 27% and 28% respectively for age, RDW, lymph, D-Dimer). *Denotes hazard ratio was not significantly different from 1 (p > 0.05).

### Increasing RDW after admission is associated with greater mortality

We investigated whether changes in RDW after admission were associated with mortality, both for those with initially elevated RDW and those without. Figure 3 shows that patients with admission RDW <= 14.5% who did not survive had an increasing RDW on average, while those with admission RDW <= 14.5% who were alive at discharge had stable RDW. For all patients, an RDW increase during admission was associated with increased mortality.

**Figure 3.**
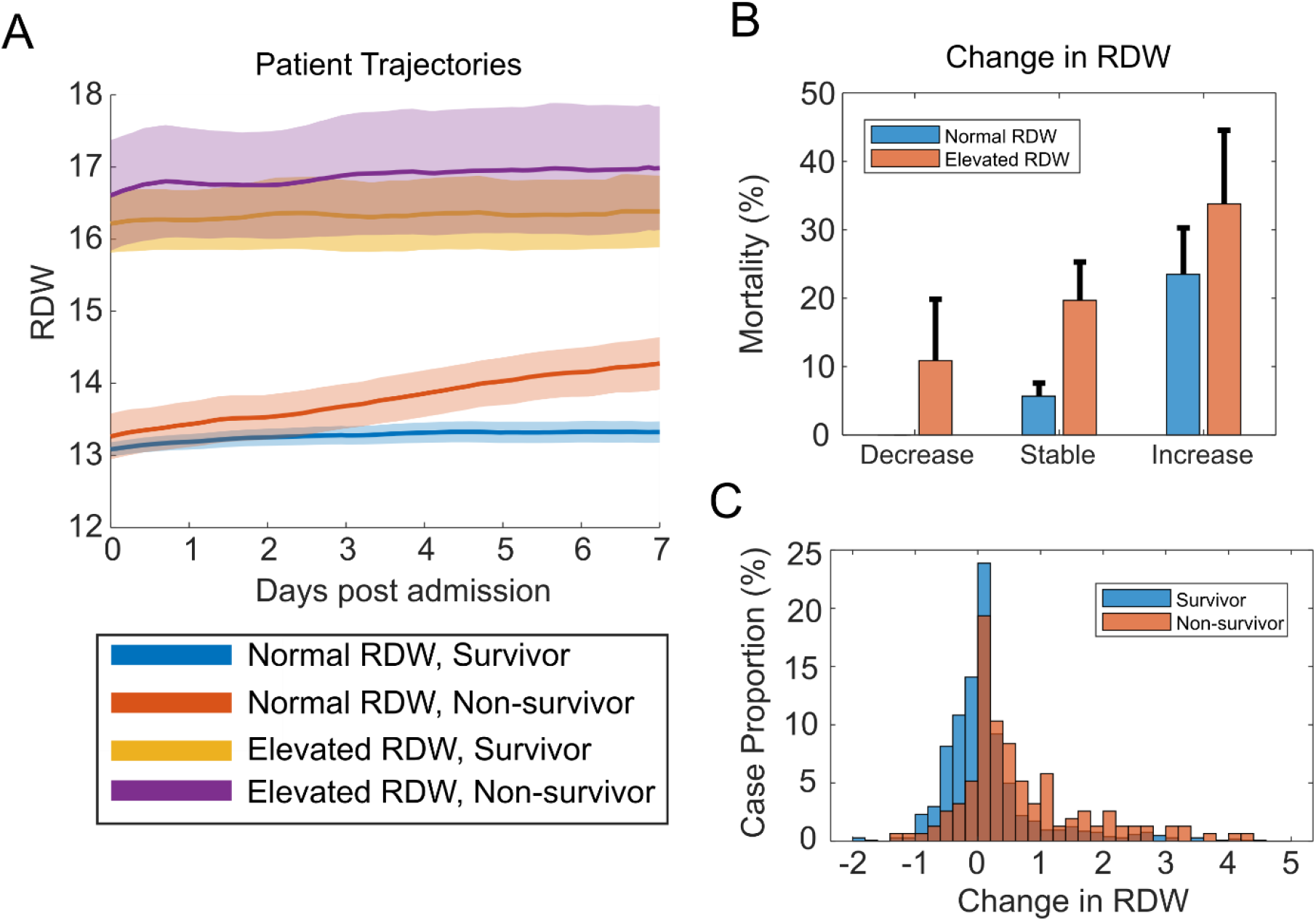
**RDW increase after admission is associated with elevated mortality risk**. (A) Stratifying patients based on admission RDW and mortality reveals that among patients with RDW <= 14.5 at admission, those who do not survive have an average RDW increase of ~1.5% during their first week of hospitalization, significantly larger than all other groups. Shading shows standard error of the mean. (B) Among patients with RDW <= 14.5 at admission, those with an increase (> 0.5%) in RDW during admission had a 23% (95% CI: 17 -30) mortality rate compared to 6% (4 - 8) for those with stable RDW (< 0.5% and > -0.5%), corresponding to a 3.8x relative risk (p-value < 1e-6). Among patients with elevated RDW at admission, a further increase in RDW during admission was associated with a mortality rate of 34% (23 – 45) and a stably elevated RDW with 20% (14 – 25), yielding a relative risk of 1.7 (p-value = 0.02). (C) A histogram of RDW change in survivors and non-survivors shows that non-survivors were more likely to experience an RDW increase during hospitalization.

## Discussion

We find that an RDW above 14.5% at the time of admission for SARS-CoV-2 infection is associated with a 2.5-fold increased risk of mortality in a cohort of 1198 adult patients treated at a large academic medical center network. RDW-associated risk of mortality remained significant after adjusting for patient age, D-Dimer, absolute lymphocyte count, and five major comorbidities. Patients whose RDW increased during admission also had an elevated mortality risk. RDW is routinely measured and may thus be helpful for prioritizing patients for earlier aggressive intervention or helping predict hospital resource usage.

The RDW-associated relative risk for younger patients (< 70) is surprisingly high with mortality rates diverging very soon after admission to reach a relative risk of 4.3 at 48 hours. This rapid decompensation is consistent with hospital presentation after significant disease progression. For patients 70-80 years, mortality rates do not diverge significantly until about 5 days after admission. Data is not available in this study to determine whether patients in this age cohort are systematically coming to the hospital earlier in the course of disease.

The specific mechanism or mechanisms for the RDW alteration in association with COVID-19 remain unclear. While RDW elevation may reflect contributions from both a decrease in MCV and an increase in RBC volume variance, the large difference in mean RDW between the elevated and non-elevated groups (~3% in Figure 3A) cannot be explain solely by the MCV difference of < 2 fL found in our cohort (see **supplemental material**). It is therefore likely that the major contributor to RDW elevation in this cohort is an increase in RBC volume variance. RDW is a non-specific marker of general illness^8-17^ and is therefore unlikely to be causally related to COVID-19 disease progression. COVID-19 is associated with altered turnover in all WBC lineages as noted above as well as with altered platelet dynamics in COVID-associated coagulopathy.^19^ The association of elevated RDW with COVID-19 severity could be consistent with prior reports suggesting that RDW can become elevated when RBC production kinetics have slowed in the setting of increased WBC and platelet kinetics.^6,8,18^

It is unknown whether patients admitted with RDW > 14.5% had higher baseline RDW than those admitted with RDW <= 14.5% prior to SARS-CoV-2 infection. RDW usually changes slowly because it reflects the volume variance of a cell population that is turning over at a rate typically no larger than a percent or two per day. Few patients in our cohort experience a > 2%/week increase in RDW during their admission, and the large increase in the elevated RDW group (> 3% on average in Figure 3A) may suggest a longer duration of disease for these patients at the time of admission, but direct study of the earlier phases of the disease is required to know how quickly RDW may be evolving prior to hospitalization. Patients with many different underlying acute and chronic illnesses would be expected to have higher baseline RDW, and it is possible that RDW at admission is providing a non-specific summary marker of the presence of these illnesses that have been shown to be associated with elevated RDW and may also be expected to complicate COVID-19 clinical course. Independent of the reasons for the differences in RDW at admission, RDW’s prognostic utility appears to persist after admission as demonstrated by the higher mortality for patients in our cohort whose RDW increased during admission.

## Data Availability

Summary data related to all figures and tables is available upon reasonable request to the author. Due to the IRB, individual patient data cannot be provided.

## Acknowledgements

This study benefited from data support provided by the Partners Healthcare Research Patient Data Registry and Enterprise Data Warehouse. This project was supported by a grant from the One Brave Idea Initiative to JMH. ADA and MBW acknowledge grant support from the CRICO Risk Management Foundation.

## References

1. Wang D, Hu B, Hu C, et al. Clinical Characteristics of 138 Hospitalized Patients With 2019 Novel Coronavirus-Infected Pneumonia in Wuhan, China. Jama 2020.

2. Grasselli G, Zangrillo A, Zanella A, et al. Baseline Characteristics and Outcomes of 1591 Patients Infected With SARS-CoV-2 Admitted to ICUs of the Lombardy Region, Italy. Jama 2020.

3. Guan WJ, Ni ZY, Hu Y, et al. Clinical Characteristics of Coronavirus Disease 2019 in China. N Engl J Med 2020.

4. Zhou F, Yu T, Du R, et al. Clinical course and risk factors for mortality of adult inpatients with COVID-19 in Wuhan, China: a retrospective cohort study. Lancet (London, England) 2020;395:1054–62.

5. Malka R, Delgado FF, Manalis SR, Higgins JM. In Vivo Volume and Hemoglobin Dynamics of Human Red Blood Cells. PLoS Computational Biology 2014;10:e1003839.

6. Higgins JM, Mahadevan L. Physiological and pathological population dynamics of circulating human red blood cells. Proc Natl Acad Sci U S A 2010;107:20587–92.

7. Cohen RM, Franco RS, Khera PK, et al. Red cell life span heterogeneity in hematologically normal people is sufficient to alter HbA1c. Blood 2008;112:4284–91.

8. Patel HH, Patel HR, Higgins JM. Modulation of red blood cell population dynamics is a fundamental homeostatic response to disease. Am J Hematol 2015;90:422–8.

9. Anderson JL, Ronnow BS, Horne BD, et al. Usefulness of a complete blood count-derived risk score to predict incident mortality in patients with suspected cardiovascular disease. Am J Cardiol 2007;99:169–74.

10. Felker GM, Allen LA, Pocock SJ, et al. Red cell distribution width as a novel prognostic marker in heart failure: data from the CHARM Program and the Duke Databank. J Am Coll Cardiol 2007;50:40–7.

11. Perlstein TS, Weuve J, Pfeffer MA, Beckman JA. Red blood cell distribution width and mortality risk in a community-based prospective cohort. Arch Intern Med 2009;169:588–94.

12. Topaz G, Kitay-Cohen Y, Peled L, et al. The association between red cell distribution width and poor outcomes in hospitalized patients with influenza. Journal of critical care 2017;41:166–9.

13. Karagoz E, Ulcay A, Tanoglu A, et al. Clinical usefulness of mean platelet volume and red blood cell distribution width to platelet ratio for predicting the severity of hepatic fibrosis in chronic hepatitis B virus patients. European journal of gastroenterology & hepatology 2014;26:1320–4.

14. Patel KV, Ferrucci L, Ershler WB, Longo DL, Guralnik JM. Red Blood Cell Distribution Width and the Risk of Death in Middle-aged and Older Adults. Archives of Internal Medicine 2009;169:515–23.

15. Perlstein TS, Weuve J, Pfeffer MA, Beckman JA. Red Blood Cell Distribution Width and Mortality Risk in a Community-Based Prospective Cohort. Archives of Internal Medicine 2009;169:588–94.

16. Patel KV, Semba RD, Ferrucci L, et al. Red Cell Distribution Width and Mortality in Older Adults: A Meta-analysis. J Gerontol Ser A-Biol Sci Med Sci 2010;65:258–65.

17. Salvagno GL, Sanchis-Gomar F, Picanza A, Lippi G. Red blood cell distribution width: A simple parameter with multiple clinical applications. Crit Rev Clin Lab Sci 2015;52:86–105.

18. Chaudhury A, Miller GD, Eichner D, Higgins JM. Single-cell modeling of routine clinical blood tests reveals transient dynamics of human response to blood loss. Elife 2019;8.

19. Spiezia L, Boscolo A, Poletto F, et al. COVID-19-Related Severe Hypercoagulability in Patients Admitted to Intensive Care Unit for Acute Respiratory Failure. Thrombosis and haemostasis 2020.

